# How does date-rounding affect phylodynamic inference for public health?

**DOI:** 10.1101/2024.09.11.24313508

**Authors:** Leo A. Featherstone, Danielle J. Ingle, Wytamma Wirth, Sebastian Duchene

## Abstract

Phylodynamic analyses enable the inference of epidemiological parameters from pathogen genome sequences for enhanced genomic surveillance in public health. Pathogen genome sequences and their associated sampling times are the essential data in every analysis. However, sampling times are usually associated with hospitalisation or testing dates and can sometimes be used to identify individual patients, posing a threat to patient confidentiality. To lower this risk, sampling times are often given with reduced date-resolution to the month or year, which can potentially bias inference of epidemiological parameters. Here, we characterise the extent to which reduced date-resolution biases phylodynamic analyses across a diverse range of empirical and simulated datasets. We develop a practical guideline on when date-rounding biases phylodynamic inference and we show that this bias is both unpredictable in its direction and compounds with decreasing date-resolution, higher substitution rates, and shorter sampling intervals. We conclude by discussing future solutions that prioritise patient confidentiality and propose a method for safer sharing of sampling dates by translating them uniformly by a random number.

## Introduction

Phylodynamics is commonly used to estimate the parameters of viral spread with increasing application to bacteria. It allows estimation of important epidemiological quantities including rates of transmission, the age of outbreaks, rates of spatial advance, and the prevalence of variants of concern (Attwood *et al*., 2022, du Plessis and Stadler, 2015, Featherstone *et al*., 2022, Volz, 2023). It is applicable across the scales of transmission from the pandemic and epidemic scales, such as for SARS-CoV-2 and Ebola virus (Lancet, 2021, Mbala-Kingebeni *et al*., 2019), to long-term bacterial transmission such as in *Salmonella enterica* and *Klebsiella pneumoniae*. Phylodynamic analyses are most useful where temporal and spatial records of transmission are sparse, using genomic information to help fill in the gaps.

The basis of all phylodynamic inference is that epidemiological spread leaves a trace in the form of substitutions in pathogen genomes that can be used to reconstruct transmission histories. Pathogen populations meeting this assumption are said to be ‘measurably evolving populations’ (Biek *et al*., 2015, Drummond *et al*., 2003). In accordance, phylodynamics uses a combination of genome sequences and associated sampling times to leverage measurable evolution and infer temporally explicit parameters of transmission and pathogen demography.

Ideal phylodynamic datasets should include precise sampling dates alongside genome sequences (Black *et al*., 2020), but sampling times necessarily carry over sensitive information about times of hospitalisation, testing, or treatment than can be used to identify individual patients. This can pose an unacceptable risk for patient confidentiality. In some cases, sampling times or dates of admission are even available for purchase or have allowed identification for a majority of patients in a given record (Sweeney, 2013). In acknowledgement of this risk, Shean and Greninger (2018) suggest that Expert Determination govern whether sampling times be released alongside genome sequences, and the resolution to which they are disclosed (day, month, year). Essentially, this approach involves an expert opinion on whether information is safe to release on a case-by-case basis.

From a phylodynamic point of view, sampling times with reduced resolution are usable. Uncertainty in sampling times can be accommodated in Bayesian inference (Shapiro *et al*., 2011), but such an approach is only effective when samples with uncertain dates comprise a small proportion of the total data (Rieux and Khatchikian, 2017).

The most common technique for incorporating data with a majority of uncertain sampling times is to assume that sampling occurred at the middle of the uncertainty range, such as all samples from 2020 being assigned 15 June 2020. Other approaches would include sampling a random day within 2020 using a probability distribution over the duration of 2020 for each sample. Both approaches introduce a degree of error, which may cause bias because sampling times can drive phylodynamic inference (Featherstone *et al*., 2021, 2023, Volz and Frost, 2014). Understanding this bias has practical significance, as there are many examples of phylodynamic analyses conducted with reduced date resolution for a diverse array of pathogens. These include viral pathogens such as Rabies Virus, Enterovirus, SARS-CoV-2, Dengue virus (Bennett *et al*., 2010, Talbi *et al*., 2010, Wolf *et al*., 2022, Xiao *et al*., 2022), and bacterial pathogens, such as *Klebsiella pneumoniae*, *Streptococcus pneumoniae*, and *Mycobacterium tuberculosis* (Azarian *et al*., 2018, Cella *et al*., 2017, Merker *et al*., 2015).

Precision in sampling dates is also relevant to the design and curation of pathogen sequence databases because sampling dates are often considered as metadata, and thus recorded inconsistently throughout repositories (Raza and Luheshi, 2016). For example, as of early September 2024, there were roughly 19.9M SARS-CoV-2 genome sequences available on GISAID with roughly 2.4% (382K) of these having incomplete date information, where sampling dates are absent or only given to the month or year. In other words, roughly 1 in 50 sequences lacked clear date resolution, reflecting global inconsistency in SARS-CoV-2 sampling time records.

In recognition of this issue, we characterised the conditions under which biases arise from reduced date resolution in phylodynamic inference. We analysed four empirical datasets of SARS-CoV-2, H1N1 Influenza, *M. tuberculosis*, *Staphylococcus aureus*, and conducted a simulation study with parameters corresponding to each empirical dataset. These pathogens are key examples of candidates for genome surveillance, with SARS-CoV-2 and H1N1 having caused pandemics and *S. aureus* and *M. tuberculosis* being global priority pathogens (WHO, 2024). These data also have diverse infectious periods and molecular evolutionary rates, thus providing a broad representation of phylodynamics’ applicability to pathogens presenting human-health threats. For each empirical and simulated dataset, we studied the bias in estimated epidemiological parameters across treatments with sampling times rounded to the day, month, or year. For example, 2021-10-11 would be specified as 2021-10-15 when rounding to the month and 2021-06-15 when the month and day are not provided.

We focused on inference of the reproductive number (*R*_0_ or *R_e_* for the basic and effective reproductive number, respectively), defined as the average number of secondary infections stemming from an individual case (reviewed by du Plessis and Stadler (2015), Featherstone *et al*. (2022), Kühnert *et al*. (2011)), the time to the most recent common ancestor (tMRCA), and the substitution rate (substitutions per site per year) in each dataset. Together, these parameters span much of the insight that phylodynamics offers through inferring when an outbreak started and how fast it proceeded. The evolutionary rate is also the central parameter relating evolutionary time to epidemiological time, so any resulting bias in this parameter is expected to have a pervasive effect throughout each phylodynamic model.

We hypothesised that reduced date resolution causes bias that compounds where the uncertainty in dates exceeds the average time for a substitution to arise in a given pathogen. We visualise the relationship between date resolution and average substitution time in Fig 1. For example, H1N1 influenza virus accumulates substitutions at a rate of about 4 ×10*^−^*^3^ subs/site/year (Hedge *et al*., 2013). With a genome length of 13,158bp, we then expect roughly one substitution to accrue per week. Therefore, rounding dates to the month or year conflates molecular evolution in time and biases inferences. Based on this, we expected the SARS-CoV-2 and H1N1 datasets to exhibit bias from month resolution onwards, the *S. aureus* dataset to exhibit bias at year resolution, and the *M. tuberculosis* dataset to not display bias up to and including year resolution (See Table 2 for average substitution times).

**Figure 1:**
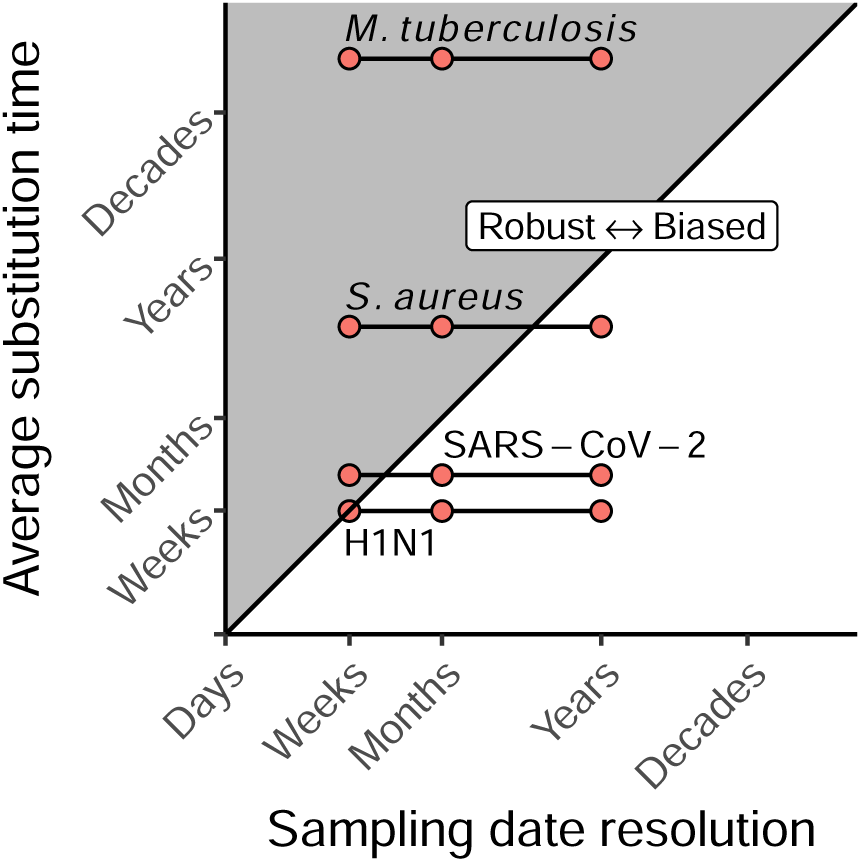
The average time to accrue one substitution based on a fixed genome size and evolutionary rate, *T_s_*= [Genome Length (sites) × Evolutionary rate (subs/site/yr)]*^−^*^1^ against the temporal resolution lost by date-rounding. We hypothesised and showed that when analyses for a given pathogen round dates to an extent nearing or crossing the diagonal from left to right, biases is induced in *R_e_*, tMRCA, and substitution rate. substitution rates are taken from each source for the empirical data. We do not report the numerical axis as this figure is designed to illustrate a concept rather than serve as a reference, in the same spirit as is inspiration in Figure 2 of Biek *et al*. (2015).

Our results across the simulation study and analyses of empirical data support using the average substitution time as a rough threshold for when date-rounding causes compounding bias. We also discuss factors that modulate the extent of bias, such as duration of sampling intervals and the choice of phylodynamic model. We finish by discussing future solutions that prioritise both patient confidentiality and accurate data sharing for routine phylodynamic analyses for public health.

## Methods

### Overview

Our study is based on four empirical datasets including with two viruses, H1N1 influenza and SARS-CoV-2, and two bacterial species, *Staphylococcus aureus* and *Mycobacterium tuberculosis*. We also conducted a simulation study with parameters tailored to each dataset. These data were chosen to span the usual parameter space for substitution rate and sampling duration in phylodynamics for epidemiology (roughly 10*^−^*^3^-to-10*^−^*^8^ (subs/site/yr) for substitution rate and months-to-decades for duration of sampling).

To assess the effects of date-rounding, we conducted phylodynamic analyses for both the empirical and simulated datasets with sampling dates rounded to the day, month, or year. For example, two samples from 2000-05-29 and 2000-05-02 would become 2000-05-15 if rounded to the month. We then measured the resulting bias in epidemiologically- or phylodynamically-important parameters: the reproductive number (*R*_0_ or *R_e_*), substitution rate (subs/site/year), and the tMRCA. The tMRCA gives a measure of the age of the pathogen population driving the outbreak and is often interpreted as the age of the outbreak. We also consider the tMRCA to facilitate comparison, because there is variability in which phylodynamic models include the length of the root branch in the age of the outbreak (Stadler *et al*., 2012).

The two viral datasets consist of samples from the 2009 H1N1 pandemic (n=161) from Hedge *et al*. (2013), and a cluster of early SARS-CoV-2 cases from Victoria, Australia in 2020 (n = 112) (Lane *et al*., 2021). The bacterial datasets consist of *S. aureus*, with 104 samples from New York sampled over ≈2 years (Ducĥene *et al*., 2016, Uhlemann *et al*., 2014, Volz and Didelot, 2018), and 30 *M. tuberculosis* samples from an ≈25 year outbreak studied by Kühnert *et al*. (2018). These data were chosen because they encompass a diversity of epidemiological dynamics, timescales, and variable substitution rates.

### Simulation Study

We simulated outbreaks as birth-death sampling processes using the ReMaster package in BEAST v2.7.6 (Bouckaert *et al*., 2019, Vaughan, 2024). Simulations consisted of four parameter settings corresponding to each empirical dataset (Table 1), with 100 replicates of each. All parameter sets include a proportion of sequenced cases (*p*), outbreak duration (*T* ), and a ‘becoming un-infectious’ rate 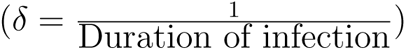. For simulations corresponding the viral datasets, transmission was modelled via *R*_0_, the average number of secondary infections (assuming a fully susceptible population). For those corresponding to the bacterial datasets, we allowed the effective reproductive numbers to vary over two intervals (*R_e_*_1_ and *R_e_*_2_ respectively). For the *S. aureus* setting, the change time for *R_e_* was set at *t* = 22 with the sequencing proportion (*p*) also set to zero before this time to replicate the sampling effort in the empirical dataset. For the *M. tuberculosis* dataset, the change time was fixed at halfway through simulations (*t* = 12.5) with a fixed sequencing proportion throughout.

**Table 1:** Parameter sets for the simulation study corresponding to each empirical dataset. *δ* is the ‘becoming un-infectious‘ rate, which is the reciprocal of the duration of infection in units of years*^−^*^1^. *R*_0_ is the basic reproductive number, describing the average number of secondary infections arising at the beginning of an outbreak where the susceptible population is greatest. *R_e•_* refer to the effective reproductive number over two successive intervals of an outbreak as the susceptible population varies. *p* is the proportion of sequenced cases. *T* is the duration of the outbreak.

| Microbe | $\delta(\text{yrs})^{-1}$ | $R_0$ | $R_{e_1}$ | $R_{e_2}$ | $p$ | $T(\text{yrs})$ | Source |
| --- | --- | --- | --- | --- | --- | --- | --- |
| H1N1 | 91.31 | 1.3 | - | - | 0.015 | 0.25 | Hedge et al. (2013) |
| SARS-CoV-2 | 36.56 | 2.5 | - | - | 0.80 | 0.16 | Lane et al. (2021) |
| *S. aureus* | 0.93 | - | 2.0 | 1.0 | 0.2<sup>†</sup> | 25 | Duchêne et al. (2016) |
| *M. tuberculosis* | 0.125 | - | 2.0 | 1.10 | 0.08 | 25.0 | Kühnert et al. (2018) |

<sup>†</sup> $p$ was set to zero before $T = 22$

Simulations generated a total of 400 outbreaks which we then used to simulate sequences data under a Jukes-Cantor model using Seq-Gen v1.3.4 (Rambaut and Grass, 1997) with fixed substitution rates (Table 2). We chose a simple substitution model to reduce parameter space and because substitution model mismatch has been widely explored elsewhere (e.g. Lemmon and Moriarty (2004)).

**Table 2:** Substitution rates and genome length for sequence simulation.

| Microbe | Substitution Rate (subs/site/yr) | Genome Length | Time/Sub/Genome (yrs) |
| --- | --- | --- | --- |
| H1N1 | $4 \times 10^{-3}$ | 13158 | 0.0190 |
| SARS-CoV-2 | $1 \times 10^{-3}$ | 29903 | 0.0334 |
| *S. aureus* | $1 \times 10^{-6}$ | 2900000 | 0.3458 |
| *M. tuberculosis* | $1 \times 10^{-7}$ | 4300000 | 2.3256 |

We then analysed each of the 400 simulated datasets under each tree prior and three date resolutions (day, month, and year), yielding 1800 analyses (1200 for the birth-death and 600 for coalescent with exponential growth, referred to hereon as the ‘coalescent exponential‘ or CE). We used identical model specifications and prior distributions as for the corresponding empirical datasets. We ran each MCMC chain for 5 × 18^8^ steps, sampling every 10^4th^ step and discarding the first 50% as burnin. We then discarded all analyses that did not have effective sample sizes of the MCMC (*ESS*) of at least 200 (*ESS* ≥ 200), leaving a total of 1670 replicates incorporated in our results.

### Empirical Data

We conducted Bayesian phylodynamic analyses using a birth-death skyline tree prior in BEAST v2.7.6 for all datasets (Bouckaert *et al*., 2019, Stadler *et al*., 2012). We also fit a coalescent tree prior with exponential population growth for the viral datasets (Kingman, 1982). We sampled from the posterior distribution using Markov chain Monte Carlo (MCMC), with 5 × 10^7^ steps (1 × 10^7^ for SARS-CoV-2 data), sampling every 10^4^ steps, and discarding the initial 10% as burnin. We assessed sufficient sampling from the stationary distribution by ensuring *ESS* ≥ 200 for all parameters and likelihoods.

### H1N1

The H1N1 data consist of 161 samples from North America during the 2009 H1N1 influenza virus pandemic, previously analysed by Hedge *et al*. (2013). Samples originate from April to September 2009 and provide an example of a rapidly evolving pathogen sparsely sequenced during an emerging outbreak.

Under the birth-death model, we placed a Lognormal(*µ* = 0*, σ* = 1) prior on *R*_0_, *β*(1, 1) prior on *p*, and fixed the becoming-uninfectious rate to (*δ* = 91 *years^−^*^1^), corresponding to a four-day duration of infection. We also placed an improper (*U* (0, ∞)) prior on the age of the outbreak and a Gamma(shape = 2, rate = 400) prior on the substitution rate.

Under the coalescent exponential, we placed a Laplace(*µ* = 0, scale = 100) prior on the growth rate, which was later transformed to *R*_0_ via *R*_0_ = *rD* + 1 where *r* is the growth rate and *D* is the duration of infection. We also placed an improper prior on the effective population size, and otherwise included the same priors as for the birth-death.

### SARS-CoV-2

The SARS-CoV-2 data consist of 112 samples from a densely sequenced transmission cluster from Victoria, Australia over late July to mid September 2020 Lane *et al*. (2021). These data are similar to the H1N1 datasets in presenting a quickly evolving viral pathogen, but differ in that a high proportion of cases were sequenced.

Under the birth-death, we placed a Lognormal(mean = 1, sd = 1.25) prior on *R*_0_ and an Inv-Gamma(*α* = 5.807*, β* = 346.020) prior on the becoming-uninfectious rate (*δ*). The sampling proportion was fixed to *p* = 0.8 since every the target was to sequence every known SARS-CoV-2 case in Victoria at this stage of the pandemic, with a roughly 20% sequencing failure rate. We also placed an Exp(mean = 0.019) prior on the origin, corresponding to a lag of up to one week between the index case and the first putative transmission event. Lastly, we placed a Gamma(shape = 2, rate = 2000) prior on the substitution rate.

Under the coalescent exponential, we placed an improper prior ( <u>^1^</u> ) on the effective population size and a Laplace(*µ* = 0.01, scale = 0.5) prior on the growth rate. Other parameters were given the priors as under the birth-death. Note that we fit the coalescent exponential tree prior for completeness here, but in practice it would not be a reliable model choice due to the high sequencing proportion violating the assumption of a low sequencing proportion under the coalescent. This poor fit is reflected later in the results.

### Staphylococcus aureus

The *S. aureus* dataset originates from Uhlemann *et al*. (2014) and we analysed a subset of the data later analysed in Ducĥene *et al*. (2016) and Volz and Didelot (2018). It consists of a single nucleotide polymorphism (SNP) alignment of 104 sequenced isolates sampled in New York from 2009 to 2011. Populations growth is understood to have been driven by *β*-lactam antibiotic use beginning in the 1980s. These data therefore provide a comparison to the *M. tuberculosis* dataset in a briefer sampling span from an outbreak of similar duration.

To accommodate changing transmission dynamics, we included two intervals for *R_e_* with a Lognormal(*µ* = 0*, σ* = 1) prior on each. We also placed a *β*(1, 1) prior on the sampling proportion, which was otherwise fixed to 0 before the first sample to capture the lag in sampling. We also placed a *U* (0, 1000) prior on the origin, and fixed the becoming un-infectious rate at *δ* = 0.93, corresponding to a nearly year-long duration of infection following Volz and Didelot (2018).

### Mycobacterium tuberculosis

The *M. tuberculosis* dataset consists of 36 sequenced isolates from a retrospectively recognised outbreak in California, USA, that originated in the Wat Tham Krabok refugee camp in Thailand. The data were originally analysed using the birth-tree prior by Kühnert *et al*. (2018). We applied the same prior configurations as Kühnert *et al*. (2018), with the exception of including two intervals for *R_e_* and fitting a strict molecular clock with a Gamma(shape = 0.001, rate = 1000.0) prior.

## Results

### Simulation study

The viral simulation conditions (i.e. SARS-CoV-2 and H1N1) display the greatest bias in mean posterior estimates of substitution rate, tMRCA, and reproductive number with decreasing date resolution (Figure 2 A-C). The *S. aureus* simulations exhibit similar trend with lesser bias in response to decreasing date resolution when rounding dates to the year. The *M. tuberculosis* condition is effectively inert to decreasing date resolution, with mean posterior estimates for each parameter of interest remaining consistent across date resolution (day to year). The *S. aureus* data provide an important intermediate case in that estimates of each parameter change when transitioning from month to year resolution (see crossing of lines from month to year resolution in the *S. aureus* column of Figure 2). These trends are in agreement with the hypothesis of decreasing date resolution causing increased bias where the resolution lost exceeds the average time for a substitution to arise. This occurs because date-rounding compresses divergent sequences in time, driving a signal for higher rates of substitution and transmission locally to each temporal cluster of sampling times. This effect is less pronounced in the bacterial simulation conditions relative to the viral conditions, because the date resolution lost is a smaller fraction of the effective substitution time (average time to until substitution is ≈ 4 months and ≈ 28 months for *S. aureus* and *M. tuberculosis* conditions respectively, Table 2). In other words, the bacterial sequences clustered in time were on average less divergent than for the viral data, which is biologically realistic given that bacteria tend to accrue substitutions more slowly than viruses. There are also notable deviations from these general trends across date resolutions, simulation conditions, and tree priors that we attribute to the duration of the sampling intervals below.

**Figure 2:**
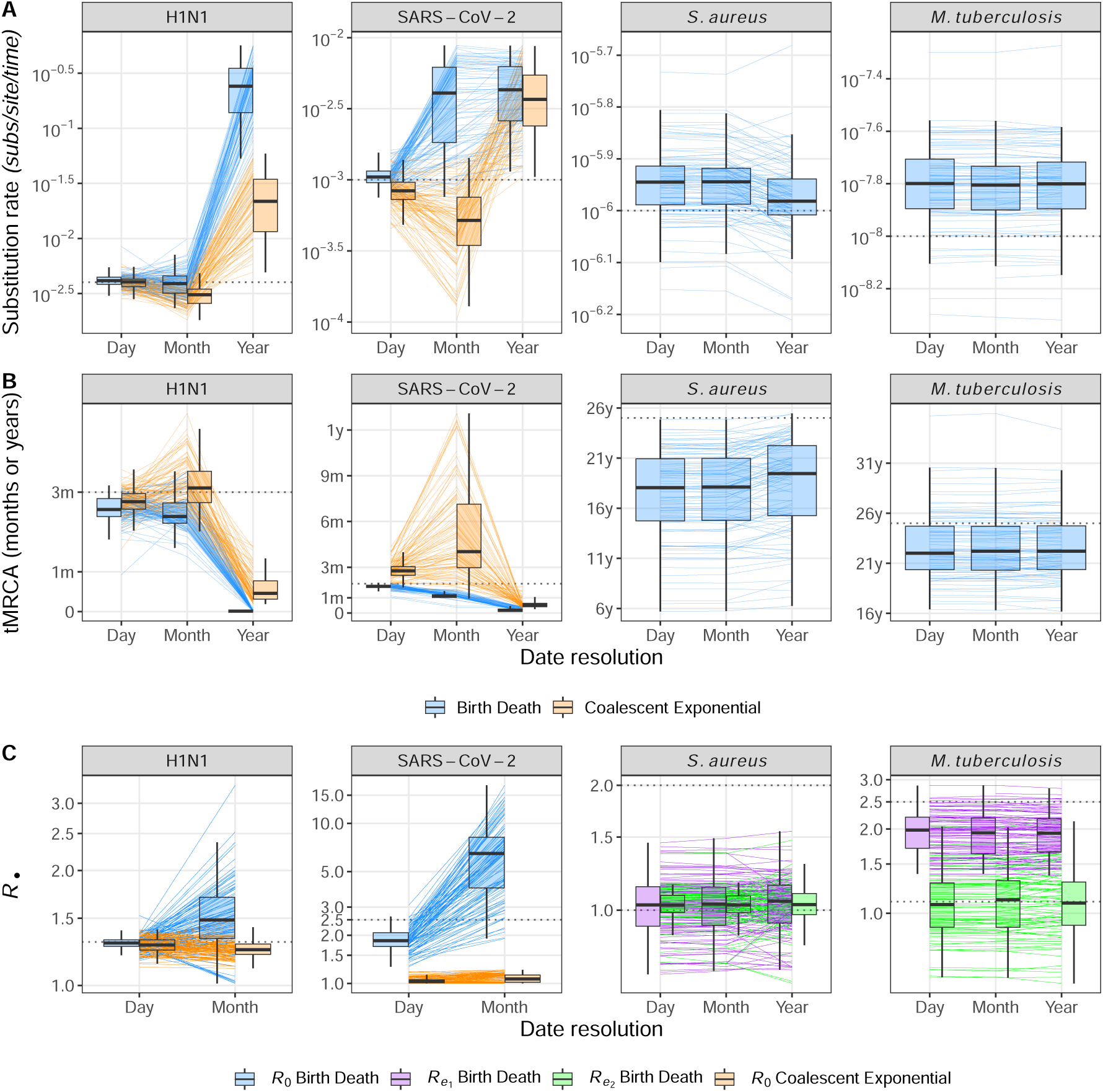
Mean posterior estimates for parameters of interest for each simulated dataset varying across date resolution. Individual lines track mean posterior estimates for each simulated dataset and boxplots are given to summarise the spread and direction of bias across all simulated datasets at each date resolution. Rows correspond to individual parameters, columns correspond to simulation conditions (underlying parameters matching each empirical dataset), and colour corresponds to tree prior or reproductive number interval. Dashed horizontal lines correspond to the true value under which each dataset was simulated. (**A**) Mean posterior substitution rate across simulation scenarios. (**B**) Mean posterior tMRCA, a measure of the age of the population driving the outbreak. (**C**) mean posterior reproductive number.

The coalescent exponential shows overall downwards bias in the substitution rate for the SARS-CoV-2 and H1N1 treatments at month resolution, while the birth-death exhibits upwards bias. Since the sampling times for each viral dataset are distributed over three months, date-rounding compresses samples within a month to one time, simultaneously increasing the time between samples across months and driving a signal for lower transmission and substitution rates between months. The different phylodynamic likelihood functions for each tree prior respond differently to this warped distribution of diversity over time with the coalescent exponential placing more weight on decreased rates of substitution rates while the birth-death favoured an increase. This can be explained by the birth-death drawing signal for increased transmission among coincident sampling times within each month, while the coalescent exponential instead conditions on sampling times (Volz and Frost, 2014). At the year resolution there is there is also lower bias in estimates of substitution rate for the coalescent exponential than the birth-death, however both models estimate upwards-biased substitution rates as year resolution. This is probably because year resolution clusters all sampling times to a single time, meaning a highly inflated rate of substitution is needed to model the artificial burst in diversity at one time for both tree priors - See Figure S2 to see sampling times compressed in time across date resolution for posterior trees. For all viral simulation conditions, the mean posterior tMRCA of each outbreak shifts inversely to the substitution rate. This is the result of a well understood relationship among phylodynamic models where higher rates of evolution suggest shorter periods of evolution.

The reproductive number for each viral dataset (*R*_0_) also changes markedly with decreasing date resolution under the birth-death, but not under the coalescent. For the birth-death, this is in agreement with temporal clustering of samples driving a signal for higher transmission rates. Conversely, estimates under the coalescent exponential remain near-identical at month resolution, which is again due to its conditioning on sampling times. Estimates of *R*_0_ for the SARS-CoV-2 settings under the coalescent exponential are also heavily biased downwards. This is probably due to high sequencing proportions violating the assumption of low sampling under the coalescent, thus leading to poorly fitting model in the first place.

The *S. aureus* condition yields consistent estimates of substitution rate, tMRCA, and reproductive number (*R_e_* in this case) when days are rounded to the month (Figure 2 *S. aureus* column). At year resolution the posterior substitution rate appears biased downwards. This can be explained by the two year sampling duration of the *S. aureus* condition, such that samples rounded to the year will be on average further apart in time than if dates are given to the month or day (Figure S2). This spacing of diversity in time likely drives the signal for lower substitution rates and an older outbreak in turn. There is no clear pattern in the direction of bias for *R_e_*_1_ and *R_e_*_2_ at year resolution, though estimates deviate from those at month and day resolution. Estimates for *R_e_*_1_ are also overall lower than their true value of 2.0, and this is attributable to inconsistent sampling over the duration of the outbreak which was previously demonstrated for other datasets with late sampling in Featherstone *et al*. (2021).

The *M. tuberculosis* simulation condition effectively acts as a control, since it appears inert to date-rounding. This is expected because this dataset reflects longer simulation time, with temporal clustering less likely to inflate *R_e_*, and an average substitution time is longer than a year. As such, even rounding to the year is unlikely to drive a signal for increased evolutionary rate or a more recent origin time.

Phylodynamic and phylogenetic terms from the total posterior likelihood also vary with decreasing date resolution (Figure S3). deviation also increases with lesser date resolution from month and year. This verifies that altered date resolution affects the likelihood manifold of each analysis, which is reflected in the different trends of bias in each parameter of interest.

### Empirical Results

Broadly, analyses of the empirical datasets reproduce the patterns of bias in the simulation study(Figure 3). That is, the reproductive number increases with decreasing date resolution along with an increase in the substitution rate and corresponding decrease in the tMRCA. There are a few exceptions to this trend that we consider below and which we again attribute to the difference between simulated and empirical sampling time distributions.

**Figure 3:**
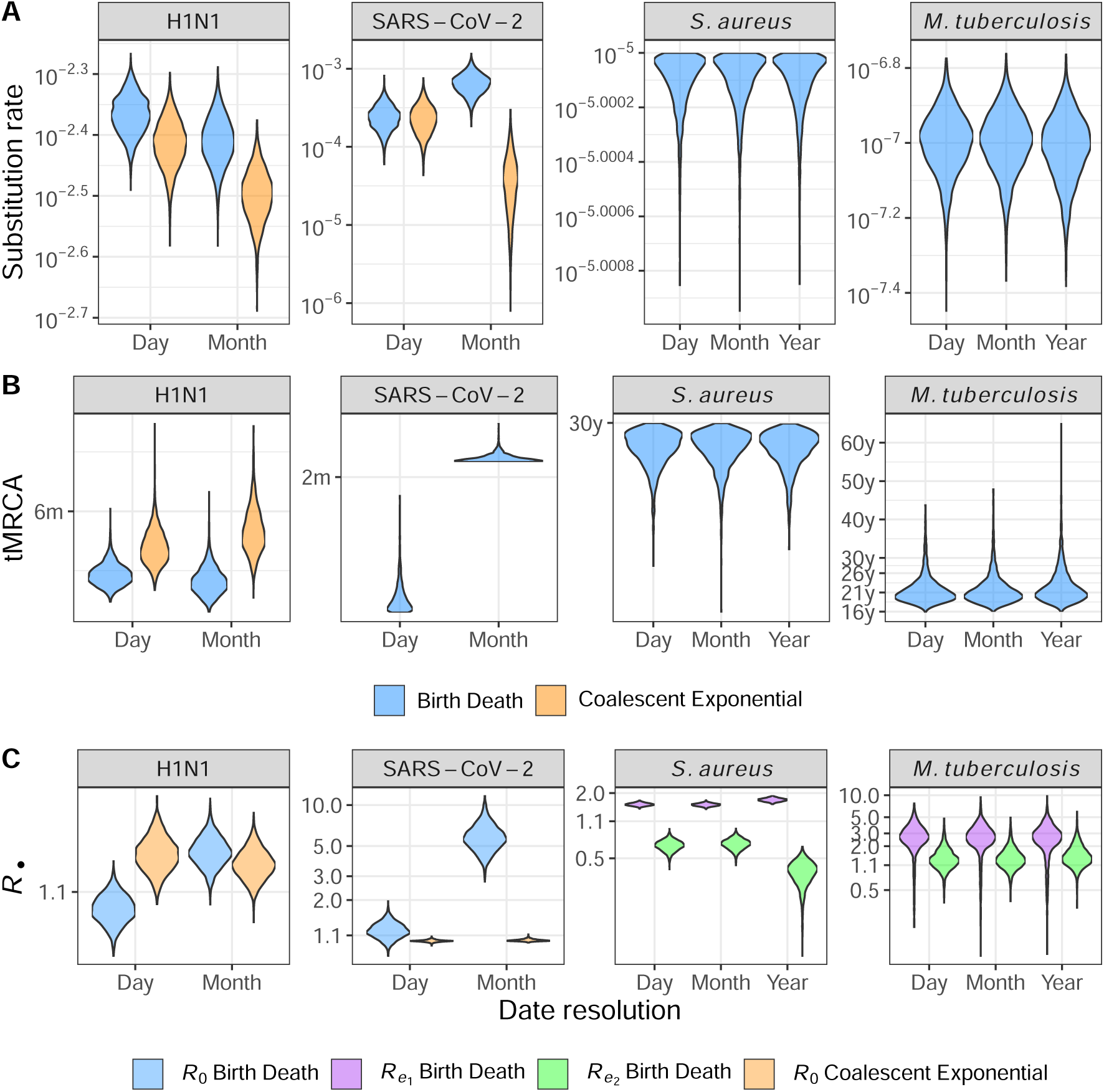
Posterior distributions for parameters of interest estimated for each empirical dataset. Date resolution is given on the horizontal axis and colour denotes tree prior. Estimates for viral datasets at year-resolution are omitted because results deviate by implausible orders of magnitude due to sampling times rounded to identical dates. (**A**) Posterior substitution rate across date resolutions. (**B**) Posterior tMRCA in units of months (m) or years (y). (**C**) Posterior reproductive number on a log-transformed axis.

Phylodynamic and phylogenetic likelihoods also diverge where the loss in date resolution exceed the average time for a substitution to arise (Figure 4). For both viral datasets, month and day posterior distributions of phylodynamic and phylogenetic likelihood are diverged, while likelihoods overlap at all date resolutions for the *M. tuberculosis* data. The *S. aureus* data provide an intermediate case where only the posterior likelihoods for year-resolution differ. Together, these likelihood distributions support the hypothesis that date-uncertainty that is wider (in time) than the average time to one substitution causes a qualitative shift in the likelihood manifold for analyses under both birth-death based and coalescent tree priors.

**Figure 4:**
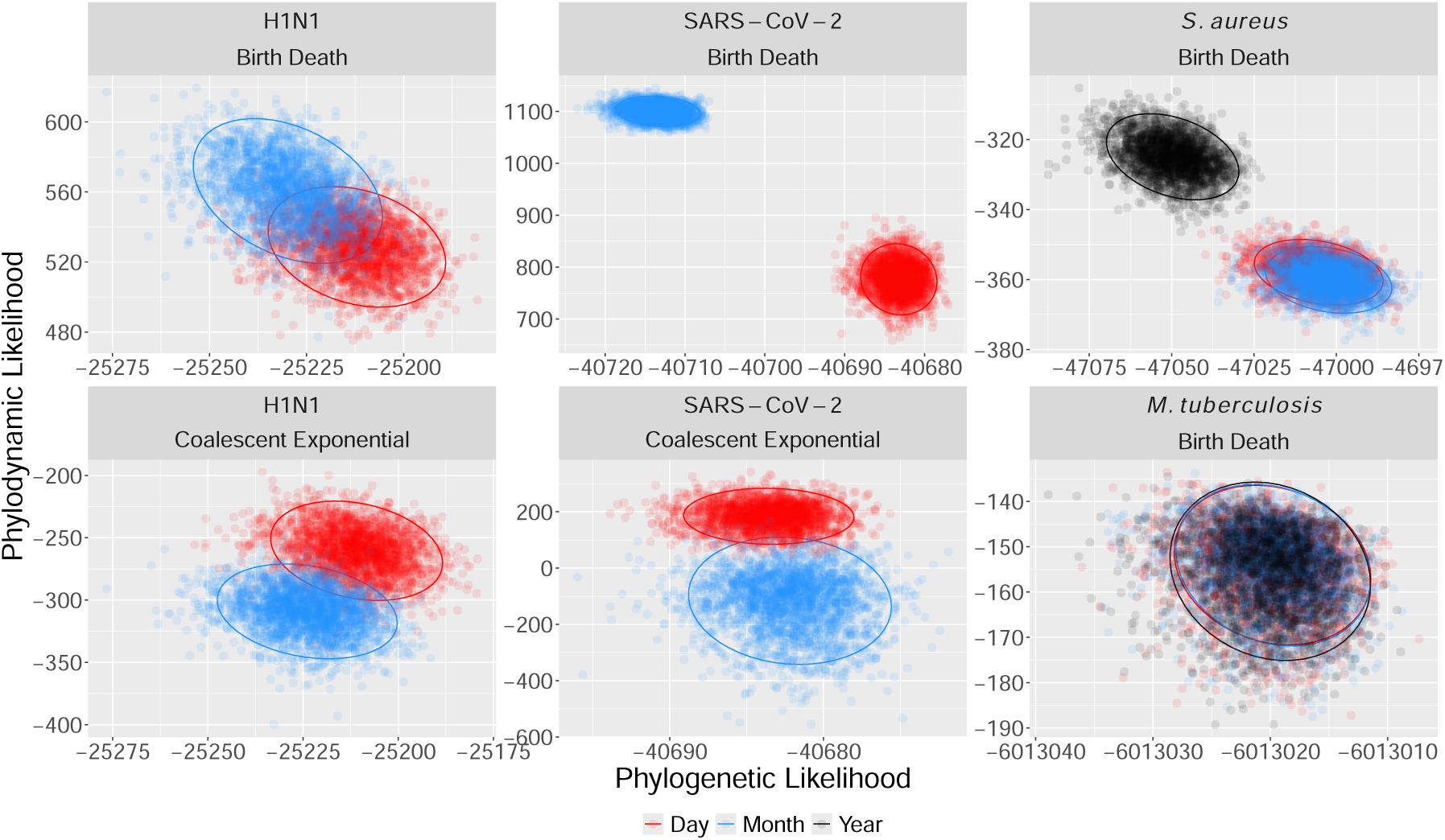
Posterior phylodynamic likelihood against phylogenetic likelihood for each combination of empirical dataset and with colour corresponding to date resolution. Ellipses surround the 95% highest posterior density region for each posterior. Both Phylodynamic and phylogenetic likelihoods diverge between day and month resolution for the viral datasets, while year resolution differs from month and day for the *S. aureus* data. Posterior likelihoods all coincide for *M. tuberculosis*.

### H1N1

Mean posterior *R*_0_ increases from day to month resolution for the birth-death (1.08 to 1.14), yet remains near-identical for the coalescent exponential (1.14 to 1.13) (Table S1). The mean posterior substitution rate also decreases for both tree priors across day to month resolution (4.3 × 10*^−^*^3^ to 3.9 × 10*^−^*^3^ and 3.9 × 10*^−^*^3^ to 3.2 × 10*^−^*^3^ for the birth-death and coalescent exponential respectively) (Table S1). The posterior tM-RCA also differs between tree priors mirroring substitution rate, with a decrease from date to month resolution for the birth-death and an increase for the coalescent exponential. For the coalescent exponential, we can attribute the decrease in reproductive number and substitution rate from day to month resolution to samples being spread further in time (Figure S1), which drives the signal for and an older outbreak and lower transmission rates. While the same is true for the sampling distribution under the birth-death, the additional information it draws from identical sampling times as month-resolution likely inflates the mean posterior reproductive number despite the signal for a lower substitution rate and older outbreak.

### SARS-CoV-2

Under the birth-death, the SARS-CoV-2 dataset behaves as expected, with an increase in posterior *R*_0_ from day to month rounding. In particular, rounding to the month results in a high, yet plausible value of *R*_0_ = 5.972 (Table S1). Under the coalescent exponential, the mean posterior *R*_0_ remains near identical across day to month treatments (1.00 to 1.01 respectively). We again note that the coalescent exponential is included for completeness for the SARS-CoV-2 dataset, but is not an appropriate choice of model in practice due to the near complete-sequencing of the original transmission cluster. Thus, poor model-fit is probably the cause of unrealistic estimates of *R*_0_.

The mean posterior substitution rate under the birth-death increases over two-fold when rounding to the month (2.47 × 10*^−^*^4^ to 6.56 × 10*^−^*^4^, Table S1). Mean posterior tMRCA also increases from 0.15 years to 0.17 years from day to month, which contradicts the expectation of a decreased estimate of tMRCA under daterounding. We again attribute these differences to the distribution of the empirical sampling times under date-rounding. Sampling for the SARS-CoV-2 dataset mainly occurred over August to September 2020, with most August samples originating later in the month (Figure S1). Rounding to 15*^th^* of August therefore made these samples appear older in time and likely contributed to the older origin under month-rounding.

### S. aureus

For *R_e_*_1_, the *S. aureus* dataset recapitulated the simulation study with month rounding having a minimal effect, but year rounding inducing an upwards bias (mean values of 1.57, 1.56, 1.73 respectively)(Figure 3, Table S1). *R_e_*_2_ displays a similar pattern with consistent estimates at day and month-rounding before a reduction at year rounding (0.66, 0.67, and 0.37 respectively). This result is consistent with the estimates an initial increase in growth rate in previous analyses of the dataset Volz and Didelot (2018).

Mean posterior substitution rate and tMRCA remain identical across date resolutions (10*^−^*^5^ subs/site/year and a tMRCA of 30 years), despite the change in reproductive numbers at year rounding. This is surprising given the change in posterior phylodynamic and phylogenetic likelihoods (Figure 4), and highlights that date-rounding can perturb the likelihood without predictable changes in parameters of epidemiological significance.

### M. tuberculosis

The *M. tuberculosis* data recapitulate the outcome of the simulation study in being robust to date-rounding. Posterior substitution rates and outbreak ages remain consistent across decreasing date resolution (1.02 × 10*^−^*^7^, 1.02 × 10*^−^*^7^, 9.86 × 10*^−^*^8^ (subs/site/time) and 21.7, 21.7, and 22.5 years respectively) (Table S1, Figure 3). We also infer that *R_e_*_1_ *> R_es_* across date-rounding conditions, coinciding with an earlier burst of transmission in agreement with Kühnert *et al*. (2018). However, *R_e_*_1_ decreases slightly date date-rounding (mean posterior estimates of 2.77, 2.74, 2.66 for day, month and year rounding)(Table S1), while *R_e_*_2_ increases (1.4, 1.41, 1.53 from day to year rounding). This was likely caused by the higher number of samples in the second sampling interval, from roughly 2002 to 2010, such that compressing sampling times drive drove a signal for higher transmission in the second interval with longer periods between sampling in the first interval at year resolution. Again, this shows that distribution of sampling times for empirical data, which is largely unpredictable, modulate the effects of date-rounding.

## Discussion

The results of the simulation study and analyses of empirical data support our hypothesis that phylodynamic inference is most biased where the temporal resolution lost in date rounding exceeds the average time for one substitution to arise. In the both the simulation study and empirical analyses, the viral datasets (H1N1 and SARS-CoV-2) display the greatest bias in mean posterior reproductive number, substitution rate, and tMRCA when rounding to the month or year, with the average substitution time being less than one month in both simulation conditions. The *S. aureus* data provide an intermediate case, with estimated parameters displaying bias when rounding dates to the year (average substitution time between the order of months to a year). Lastly, the *M. tuberculosis* data also provide supporting evidence in not displaying any notable bias between estimates at day, month, or year date-rounding. This is expected because the average substitution time longer than a year in all *M. tuberculosis* analyses.

We therefore propose the average substitution time as a rough practical threshold after which genomic epidemiologists can invariably expect date-rounding to distort inference. Genomic epidemiologists can make this assessment by calculating the average substitution time, *T_s_*, as *T_s_* = [Genome Length (sites)×Evolutionary rate (subs/site/yr)]*^−^*^1^ and checking whether 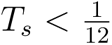 (indicating substitutions arising faster than monthly) when justifying rounding to the day, or *T_s_ <* 1 (substitutions arising more than yearly) when justifying rounding to the month. In the more general terms, we propose that date rounding is problematic for fast-evolving RNA viruses, such as in the H1N1 and SARS-CoV-2 datasets. We urge others uploading data to repositories such as GISAID to include dates to the day where possible, and support the practice of including dates to the day on pathoplexus (pathoplexus.org). This will increase the added-value of phylodynamic analysis for future infectious disease threats. Rounding to the year is sufficient for slowly evolving bacteria such as *M. tuberculosis*. We suggest case-by-case assessment for pathogens with intermediate average substitution times, such as the *S. aureus* herein and other faster-evolving bacteria including *Streptococcus*, multi-drug resistant *Escherichia coli*, or *Klebsiella pneumoniae* (Gorrie *et al*., 2018, Sherry *et al*., 2022, Xie *et al*., 2024). In the specific cases of *S. aureus* and other high disease-burden bacteria with asymptomatic and/or community carriage, we suggest preserving dates as much as possible to recover maximal information given the additional work that is often dedicated to screening samples. Finally, we note that genome samples with or without rounded dates reflect considerable efforts in the field to collect and process samples. In the case where only low-resolution dates are available, we do not discourage phylodynamic analyses, but instead encourage additional analyses to test the effects of rounded dates, such as by including priors on sampling ranges.

We also strongly emphasise that this proposal is a rough guideline lacking rigorous mathematical derivation. Any degree of date-rounding may alter likelihood and parameter estimation in phylodynamic analyses. Other factors such as as the length of the sampling window, distribution of sampling times over this interval, and choice of tree prior also affect the direction and severity of bias when rounding dates.

Shorter sampling intervals can also exacerbate the bias due to date-rounding. For example, in the SARS-CoV-2 data and simulation conditions, most sequences originated over one month with the remainder towards the end of each of the previous two months. Bias for these data was greater for each parameter compared to otherwise similar H1N1 data, which had a more even distribution of sampling over three full months. This result is in line with previous results for ancient DNA data showing that date-rounding has negligible effects for timescales of millennia or longer, which we expect to span the average substitution time several-fold (Molak *et al*., 2013). This emphasises the importance of accurate dates for phylodynamic datasets of emerging pathogens sequenced over shorter timescales, where results are likely to be the most urgent and reflect shorter sampling intervals.

The choice of tree prior also affects bias when rounding dates. For example, the coalescent exponential tended to infer decreased substitution rates while the birth-death favoured increased substitution rates across simulated and empirical viral data. The inverse trend also arose for the tMRCA. This is because the birth-death draws additional information from clustered sampling times, which serves to elevate rates of substitution and transmission, while the coalescent conditions on these and relies more on the longer duration between sampling times at month resolution for both datasets.

Taken together, the results form the simulation study and empirical data show that although date-rounding biases epidemiological estimates in a theoretically predictable directions (upwards for transmission and substitution rates, downwards for tMRCA), the intensity of the bias is difficult to predict and varies with the distribution and span of sampling times as well as tree prior. We conclude that sufficiently accurate sampling times are essential where phylodynamic insight is needed to understand infectious disease epidemiology and evolution. There does not appear to be an clear way to adjust for the bias caused otherwise. Accurate sampling times will be essential for employing phylodynamics amid future infectious disease threats.

We also acknowledge that while accurate sampling times are essential for reliable phylodynamic results, it may pose an unacceptable level of risk to patient confidentiality to release sampling times. We therefore emphasise the importance of methods that prioritise both patient confidentiality and data transparency and finish by discussing potential future solutions.

### Translating dates by random seeds

The functional component of phylodynamic data are the differences among genome sequences and among dates, rather than their absolute values. It may therefore be possible to protect patient confidentiality while sharing accurate dates by translating dates uniformly by a random number. This would protect the true sampling dates while preserving the relative times between them. For example, if the sampling times in a dataset of 3 genomes are 2000, 2001 and 2002, then data providers may randomly draw a number of 1000, which is kept secret, to shift dates. The genome-associated dates 2000, 2001 and 2002 are then shared as 3000, 3001 and 3002. While currently implausible, these translated dates are usable in phylodynamic analyses and preserve the distance between sampling times. Once results are returned the data provider can internally account for the translation in any estimated ages, such as node ages or the tMRCA, by subtracting 1000. For example if the estimated age of the outbreak (taken as the tMRCA) was 5 years before the most recent sample, then the data provider can privately estimate the outbreak’s onset as 1997 (2002 - 5), while those conducting the analysis externally can only estimate the relative age of 5 years. In the same way, intervals of transmission parameters such as *R_e_* can be placed with respect to the true sampling times. Rates, such as growth or infection rates can also be accurately estimated via this method since these are not biased by shifting dates uniformly in time.

### Distributed computing

Approaches based on distributed computing, where data are analysed across remote servers, also offer promise for maximising data transparency and patient confidentiality. For example, Santos *et al*. (2022) recently developed a method to estimate phylogenetic trees from private genome data using distributed computing and quantum crytographic protocols. Routine phylodynamic analysis for genomic surveillance may also benefit from adopting protocols from so-called swarm learning approaches that allow artificial intelligence models in precision medicine to be trained across distributed datasets (together comprising a swarm) (Warnat-Herresthal *et al*., 2021). Such approaches are in general complementary with hub-and-spoke networks, which are commonly used for storing sensitive pathogen genome data in national repositories (Hoang *et al*., 2022). We remain optimistic that future advances in distributed computing can eliminate the need for date-rounding in phylodynamic analysis.

## Data Availability

All data and code to reproduce the analyses and results are available at: https://github.com/LeoFeatherstone/pdp

## Data Availability

All analyses and data used in this work can be accessed and run as a Snakemake pipline at https://github.com/LeoFeatherstone/pdp.

## Authors’ Contributions

LAF designed the study, performed all analyses, and wrote the paper. DJI provided initial empirical data and contributed to writing of the manuscript. WW and SDG provided supervision and contributed to writing of the manuscript.

## Supplementary Material

**Figure S1:**
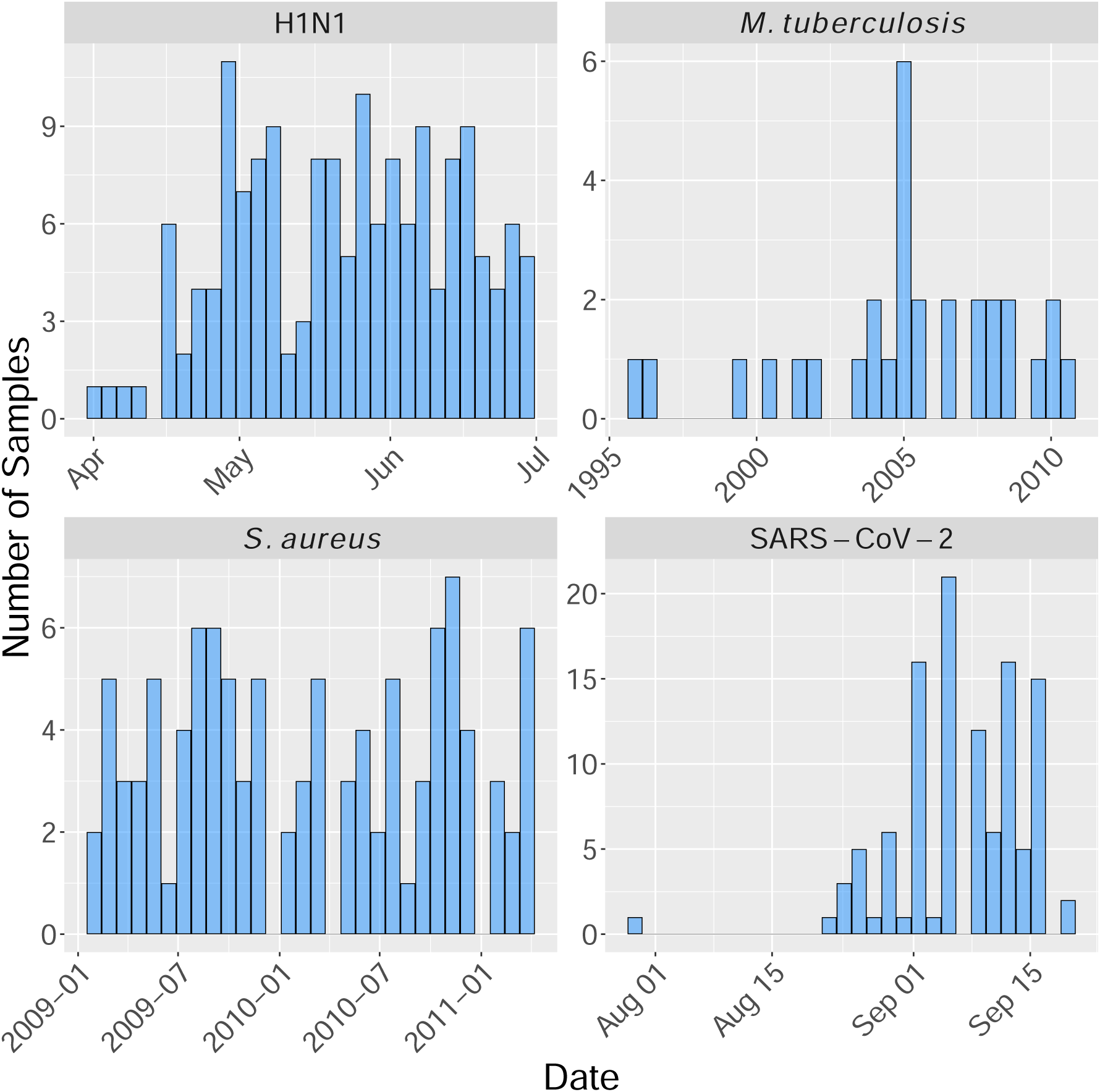
The number of samples over time for each empirical dataset. Date-rounding has the effect of moving each sampling within a month or year to the middle of that month or year (15*^th^* of the month or June 15*^th^* of the year).

**Figure S2:**
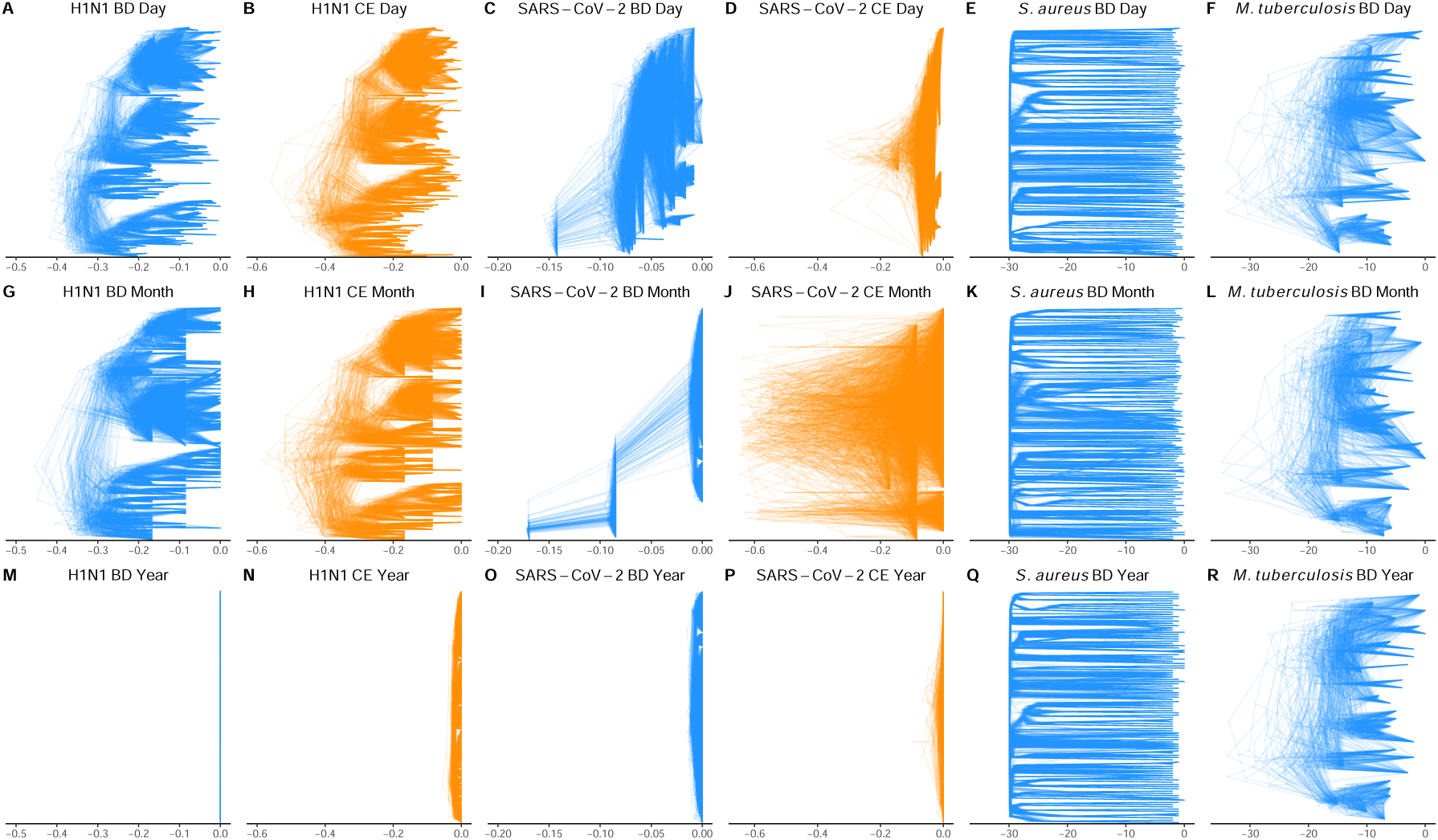
Desnsitrees (overlayed posterior trees) for empirical data with columns corresponding to pathogen under each combination of date resolution and tree prior. For the H1N1 and SARS-CoV-2 treatments, Year resolution causes trees to collapse to instantaneous bursts.

**Figure S3:**
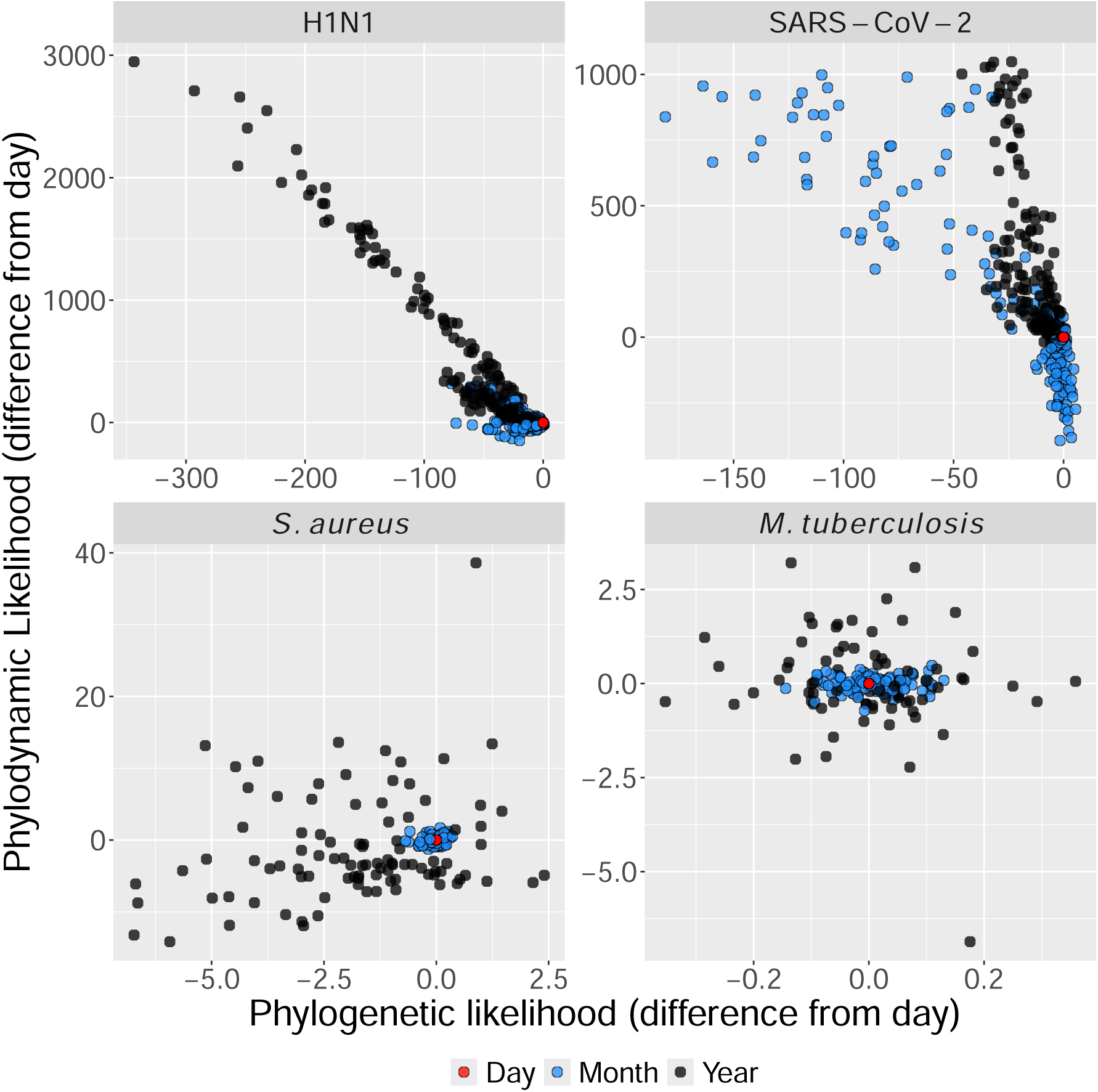
Adjusted phylodynamic likelihood against adjusted phylogenetic likelihood with panels corresponding to each simulation condition. Points correspond to mean posterior likelihood for each simulated dataset under each simulation condition. Colour corresponds to date resolution. Likelihoods are adjusted by subtracting the mean phylodynamic or phylogenetic likelihood at Day resolution from each the means under Month and year resolution. Resulting points therefore show the difference phylodynamic and phylogenetic likelihoods due to date-rounding with the point (0, 0) representing likelihood at day resolution for each dataset. Month resolution generally results in smaller differences that Year resolution, suggesting coarser date resolution results in more perturbed likelihoods. There is also generally more error in phylodynamic likelihood than phylogenetic likelihood.

**Table S1:** Mean posterior estimates of substitution rate and tMRCA for empirical data with 95% HPD in brackets. The lower table gives mean posterior estimates of *R_•_* for empirical data with 95% HPD in brackets.

|  | Tree Prior | Resolution | Substitution Rate (subs/site/yr) | tMRCA |
| --- | --- | --- | --- | --- |
| H1N1 | BD | Day | 4.31e-3 (3.7e-3, 4.9e-3) | 3.67e-1 (3.3e-1, 4.2e-1) |
| H1N1 | BD | Month | 3.9e-3 (3.2e-3, 4.6e-3) | 3.53e-1 (3.1e-1, 4.1e-1) |
| H1N1 | CE | Day | 3.87e-3 (3.2e-3, 4.5e-3) | 4.25e-1 (3.6e-1, 5.0e-1) |
| H1N1 | CE | Month | 3.17e-3 (2.6e-3, 3.8e-3) | 4.59e-1 (3.8e-1, 5.6e-1) |
| SARS-CoV-2 | BD | Day | 2.47e-4 (1.1e-4, 4.5e-4) | 1.45e-1 (1.4e-1, 1.5e-1) |
| SARS-CoV-2 | BD | Month | 6.56e-4 (3.3e-4, 1.1e-3) | 1.7e-1 (1.7e-1, 1.7e-1) |
| SARS-CoV-2 | CE | Day | 2.37e-4 (9.1e-5, 4.7e-4) | 2.03e-1 (1.4e-1, 3.6e-1) |
| SARS-CoV-2 | CE | Month | 4.34e-5 (4.4e-6, 1.4e-4) | 1.6 (2.9e-1, 5.9) |
| *S. aureus* | BD | Day | 1e-5 (1e-5, 1e-5) | 3e+01 (3e+01, 3e+01) |
| *S. aureus* | BD | Month | 1e-5 (1e-5, 1e-5) | 3e+01 (3e+01, 3e+01) |
| *S. aureus* | BD | Year | 1e-5 (1e-5, 1e-5) | 3e+01 (3e+01, 3e+01) |
| *M. tuberculosis* | BD | Day | 1.02e-7 (6.5e-8, 1.4e-7) | 2.17e+01 (1.7e+01, 3.2e+01) |
| *M. tuberculosis* | BD | Month | 1.02e-7 (6.6e-8, 1.4e-7) | 2.17e+01 (1.7e+01, 3.2e+01) |
| *M. tuberculosis* | BD | Year | 9.86e-8 (6.2e-8, 1.4e-7) | 2.25e+01 (1.8e+01, 3.4e+01) |

| | Tree Prior | Resolution | $R_0$ | $R_{e1}$ | $R_{e2}$ |
| --- | --- | --- | --- | --- | --- |
| H1N1 | BD | Day | 1.08 (1.1, 1.1) | - | - |
| H1N1 | BD | Month | 1.14 (1.1, 1.2) | - | - |
| H1N1 | CE | Day | 1.14 (1.1, 1.2) | - | - |
| H1N1 | CE | Month | 1.13 (1.1, 1.2) | - | - |
| SARS-CoV-2 | BD | Day | 1.2 (9.3e-1, 1.6) | - | - |
| SARS-CoV-2 | BD | Month | 5.85 (3.7, 9.0) | - | - |
| SARS-CoV-2 | CE | Day | 1 (9.6e-1, 1.0) | - | - |
| SARS-CoV-2 | CE | Month | 1.01 (9.8e-1, 1.1) | - | - |
| *S. aureus* | BD | Day | - | 1.57 (1.5, 1.7) | 6.56e-1 (5.1e-1, 8.0e-1) |
| *S. aureus* | BD | Month | - | 1.56 (1.5, 1.7) | 6.78e-1 (5.4e-1, 8.3e-1) |
| *S. aureus* | BD | Year | - | 1.73 (1.6, 1.8) | 3.71e-1 (1.9e-1, 5.4e-1) |
| *M. tuberculosis* | BD | Day | - | 2.77 (5.8e-1, 5.3) | 1.4 (7.2e-1, 2.7) |
| *M. tuberculosis* | BD | Month | - | 2.74 (5.7e-1, 5.0) | 1.41 (7.4e-1, 2.7) |
| *M. tuberculosis* | BD | Year | - | 2.66 (4.6e-1, 5.1) | 1.53 (8.1e-1, 2.9) |

## References

Attwood, S. W. et al. (2022). Phylogenetic and phylodynamic approaches to understanding and combating the early sars-cov-2 pandemic. Nature Reviews Genetics, 23(9), 547–562.

Azarian, T., et al. (2018). The impact of serotype-specific vaccination on phylodynamic parameters of Streptococcus pneumoniae and the pneumococcal pangenome. PLOS Pathogens, 14(4), e1006966. Publisher: Public Library of Science.

Bennett, S. et al. (2010). Epidemic Dynamics Revealed in Dengue Evolution. Molecular Biology and Evolution, 27(4), 811–818.

Biek, R. et al. (2015). Measurably evolving pathogens in the genomic era. Trends in Ecology & Evolution, 30(6), 306–313. Publisher: Elsevier.

Black, A. et al. (2020). Ten recommendations for supporting open pathogen genomic analysis in public health. Nature medicine, 26(6), 832–841.

Bouckaert, R., et al. (2019). BEAST 2.5: An advanced software platform for bayesian evolutionary analysis. PLOS Computational Biology, 15(4), e1006650. Publisher: Public Library of Science.

Cella, E. et al. (2017). Multi-drug resistant Klebsiella pneumoniae strains circulating in hospital setting: whole-genome sequencing and Bayesian phylogenetic analysis for outbreak investigations. Scientific Reports, 7(1), 3534. Number: 1 Publisher: Nature Publishing Group.

Drummond, A. J. et al. (2003). Measurably evolving populations. Trends in ecology & evolution, 18(9), 481–488.

du Plessis, L. and Stadler, T. (2015). Getting to the root of epidemic spread with phylodynamic analysis of genomic data. Trends in Microbiology, 23(7), 383–386.

Ducĥene, S., et al. (2016). Genome-scale rates of evolutionary change in bacteria. Microbial Genomics, 2(11).

Featherstone, L. A. et al. (2021). Infectious disease phylodynamics with occurrence data. Methods in Ecology and Evolution, 12(8), 1498–1507. eprint: https://onlinelibrary.wiley.com/doi/pdf/10.1111/2041-210X.13620.

Featherstone, L. A. et al. (2022). Epidemiological inference from pathogen genomes: A review of phylodynamic models and applications. Virus Evolution, 8(1), veac045.

Featherstone, L. A. et al. (2023). Decoding the Fundamental Drivers of Phylodynamic Inference. Molecular Biology and Evolution, 40(6), msad132.

Gorrie, C. L. et al. (2018). Antimicrobial-Resistant Klebsiella pneumoniae Carriage and Infection in Specialized Geriatric Care Wards Linked to Acquisition in the Referring Hospital. Clinical Infectious Diseases, 67(2), 161–170.

Hedge, J. et al. (2013). Real-time characterization of the molecular epidemiology of an influenza pandemic. Biology Letters, 9(5), 20130331.

Hoang, T. et al. (2022). AusTrakka: Fast-tracking nationalized genomics surveillance in response to the COVID-19 pandemic. Nature Communications, 13(1), 865. Publisher: Nature Publishing Group.

Kingman, J. F. C. (1982). The coalescent. Stochastic Processes and their Applications, 13(3), 235–248.

Kühnert, D., et al. (2011). Phylogenetic and epidemic modeling of rapidly evolving infectious diseases. Infection, genetics and evolution, 11(8), 1825–1841.

Kühnert, D., et al. (2018). Tuberculosis outbreak investigation using phylodynamic analysis. Epidemics, 25, 47–53.

Lancet, T. (2021). Genomic sequencing in pandemics. *Lancet (London*, England), 397(10273), 445.

Lane, C. R. et al. (2021). Genomics-informed responses in the elimination of covid-19 in victoria, australia: an observational, genomic epidemiological study. The Lancet Public Health, 6(8), e547–e556.

Lemmon, A. R. and Moriarty, E. C. (2004). The importance of proper model assumption in bayesian phylogenetics. Systematic Biology, pages 265–277.

Mbala-Kingebeni, P. et al. (2019). Medical countermeasures during the 2018 ebola virus disease outbreak in the north kivu and ituri provinces of the democratic republic of the congo: a rapid genomic assessment. The Lancet infectious diseases, 19(6), 648–657.

Merker, M. et al. (2015). Evolutionary history and global spread of the Mycobacterium tuberculosis Beijing lineage. Nature Genetics, 47(3), 242–249. Number: 3 Publisher: Nature Publishing Group.

Molak, M. et al. (2013). Phylogenetic Estimation of Timescales Using Ancient DNA: The Effects of Temporal Sampling Scheme and Uncertainty in Sample Ages. Molecular Biology and Evolution, 30(2), 253–262.

Rambaut, A. and Grass, N. C. (1997). Seq-gen: an application for the monte carlo simulation of DNA sequence evolution along phylogenetic trees. Bioinformatics, 13(3), 235–238.

Raza, S. and Luheshi, L. (2016). Big data or bust: realizing the microbial genomics revolution. Microbial Genomics, 2(2).

Rieux, A. and Khatchikian, C. E. (2017). Tipdatingbeast: An r package to assist the implementation of phylogenetic tip-dating tests using beast. Molecular Ecology Resources, 17(4), 608–613.

Santos, M. B. et al. (2022). Private Computation of Phylogenetic Trees Based on Quantum Technologies. IEEE Access, 10, 38065–38088. Conference Name: IEEE Access.

Shapiro, B. et al. (2011). A bayesian phylogenetic method to estimate unknown sequence ages. Molecular biology and evolution, 28(2), 879–887.

Shean, R. C. and Greninger, A. L. (2018). Private collection: high correlation of sample collection and patient admission date in clinical microbiological testing complicates sharing of phylodynamic metadata. Virus Evolution, 4(1), vey005.

Sherry, N. L. et al. (2022). Multi-site implementation of whole genome sequencing for hospital infection control: A prospective genomic epidemiological analysis. The Lancet Regional Health - Western Pacific, 23, 100446.

Stadler, T. et al. (2012). Estimating the basic reproductive number from viral sequence data. Molecular biology and evolution, 29(1), 347–357.

Sweeney, L. (2013). Matching Known Patients to Health Records in Washington State Data. SSRN Electronic Journal.

Talbi, C., et al. (2010). Phylodynamics and Human-Mediated Dispersal of a Zoonotic Virus. PLOS Pathogens, 6(10), e1001166. Publisher: Public Library of Science.

Uhlemann, A.-C. et al. (2014). Molecular tracing of the emergence, diversification, and transmission of S. aureus sequence type 8 in a New York community. Proceedings of the National Academy of Sciences, 111(18), 6738–6743. Publisher: Proceedings of the National Academy of Sciences.

Vaughan, T. G. (2024). ReMASTER: improved phylodynamic simulation for BEAST 2.7. Bioinformatics, 40(1), btae015.

Volz, E. (2023). Fitness, growth and transmissibility of SARS-CoV-2 genetic variants. Nature Reviews Genetics, pages 1–11. Publisher: Nature Publishing Group.

Volz, E. M. and Didelot, X. (2018). Modeling the Growth and Decline of Pathogen Effective Population Size Provides Insight into Epidemic Dynamics and Drivers of Antimicrobial Resistance. Systematic Biology, 67(4), 719–728.

Volz, E. M. and Frost, S. D. W. (2014). Sampling through time and phylodynamic inference with coalescent and birth-death models. *Journal of the Royal Society*, Interface, 11(101), 20140945.

Warnat-Herresthal, S. et al. (2021). Swarm Learning for decentralized and confidential clinical machine learning. Nature, 594(7862), 265–270. Publisher: Nature Publishing Group.

WHO (2024). WHO bacterial priority pathogens list, 2024: Bacterial pathogens of public health importance to guide research, development and strategies to prevent and control antimicrobial resistance.

Wolf, J. M. et al. (2022). Temporal spread and evolution of SARS-CoV-2 in the second pandemic wave in Brazil. Journal of Medical Virology, 94(3), 926–936. eprint: https://onlinelibrary.wiley.com/doi/pdf/10.1002/jmv.27371.

Xiao, J. et al. (2022). Genomic Epidemiology and Phylodynamic Analysis of Enterovirus A71 Reveal Its Transmission Dynamics in Asia. Microbiology Spectrum, 10(5), e01958–22. Publisher: American Society for Microbiology.

Xie, O. et al. (2024). Temporal and Geographic Strain Dynamics of Invasive Streptococcus Pyogenes In Australia: A Multi-Centre Clinical and Genomic Epidemiology Study 2011-2023. Preprints with The Lancet.

